# Calcified chondroid mesenchymal neoplasms with *FN1*-receptor tyrosine kinase gene fusions including *MERTK, TEK, FGFR2*, and *FGFR1*: a molecular and clinicopathologic analysis

**DOI:** 10.1101/2020.09.01.20186379

**Authors:** Yajuan J. Liu, Wenjing Wang, Jeffrey Yeh, Yu Wu, Jose G. Mantilla, Christopher D.M. Fletcher, Robert W. Ricciotti, Eleanor Y. Chen

## Abstract

Translocations involving *FN1* have been described in a variety of neoplasms, which share the presence of cartilage matrix and a variable extent of calcification. Fusions of *FN1* to *FGFR1* or *FGFR2* have been reported in nine soft tissue chondromas, mostly demonstrated indirectly by FISH analysis. Delineation of *FN1* fusions with various partner genes will facilitate our understanding of the pathogenesis and diagnostic classification of these neoplasms. In this study, we present molecular, clinical and pathologic features of 9 cartilaginous soft tissue neoplasms showing a predilection for the TMJ region and the extremities. We analyzed for gene fusions with precise breakpoints using targeted RNA-seq with a 115-gene panel, including *FN1, FGFR1* and *FGFR2*. All 9 cases were positive for a gene fusion, including two novel fusions, *FN1-MERTK* and *FN1-TEK*, each in one case, recurrent *FN1-FGFR2* in 5 cases, *FN1-FGFR1* without the Ig3 domain in one case, and *FGFR1-PLAG1* in one case. The breakpoints in the 5’ partner gene *FN1* ranged from exons 11-48, retaining the domains of signal peptide, FN1, FN2, and/or FN3, while the 3’partner genes retained the trans-membrane domain, tyrosine kinase domains and /or Ig domain. The tumors with *FN1-FGFR1, FN1-FGFR2* and *FN1-MERTK* fusions are generally characterized by nodular/lobular growth of polygonal to stellate cells within a chondroid matrix, often accompanied by various patterns of calcification. These features resemble those as described for the chondroblastoma-like variant of soft tissue chondroma. Additional histologic findings include calcium pyrophosphate dehydrate deposition and features resembling tenosynovial giant cell tumor. Overall, while the tumors from our series show significant morphologic overlap with chondroblastoma-like soft tissue chondroma, we describe novel findings that expand the morphologic spectrum of these neoplasms and have therefore labeled them as “calcified chondroid mesenchymal neoplasms.” These neoplasms represent a distinct pathologic entity given the presence of recurrent *FN1*-receptor tyrosine kinase fusions.

## INTRODUCTION

Translocation events involving *FN1* have been described in a wide variety of neoplasms, all of which share the presence of cartilage matrix and a variable extent of calcification (1-3). Specifically, synovial chondromatosis, characterized by multinodular growth of mature cartilaginous tissue with clustering of chondrocytes has recently been shown as frequently harboring the *FN1-ACVR2* translocation (2). Fusion of *FN1* to *FGFR1* or *FGFR2* has also been described in soft tissue chondroma, particularly in examples showing grungy to lacy (chondroblastoma-like) calcification (2). Phosphaturic mesenchymal tumor, frequently harboring *FN1-FGFR1* or *FN1-FGF1* fusions, shows variable histologic features, and is generally characterized as a proliferation of bland spindled to stellate cells with associated flocculent-appearing calcified, chondroid or ossified matrix within a highly vascularized stroma (3,4). Lastly, calcifying aponeurotic fibroma, a proliferation of bland fibroblastic cells with calcified fibrocartilage-like nodules, has been shown to harbor recurrent *FN1-EGF* gene fusions (1). Besides these four entities, these histologic features may be seen in a wide spectrum of cartilaginous neoplasms of soft tissue. These include so-called chondroid tenosynovial giant cell tumor (TGCT), calcium pyrophosphate dihydrate (CPPD) deposition disease (tophaceous pseudogout), and chondroblastoma (5-9). These tumors in some cases share overlap in cytomorphology and histologic features, sometimes making precise histologic classification challenging.

In this study, we present the molecular, clinical and pathologic features of 9 cases of soft tissue chondroid tumors with calcification, the majority of which show an anatomic predilection for the temporo-mandibular joint (TMJ). We also review the literature and discuss differential diagnosis of various entities highlighting their overlapping and distinct histologic and molecular features.

## METHODS AND MATERIALS

### Case selection and clinicopathological characterization

This project was approved by the Institutional Review Board at the University of Washington. Six cases from the series in this study were retrieved from the archives of the Department of Pathology at the University of Washington, and 3 cases were retrieved from the archived cases in consultation to C.D.F. at the Brigham and Women’s Hospital. The H&E slides were independently reviewed by 4 pathologists, E.C., R.R., J.M. and C.D.M.F.

### Targeted RNA sequencing

All nine specimens used in this study were archived formalin-fixed paraffin embedded (FFPE) tissue specimens. Total nucleic acid (TNA) was extracted from the FFPE specimens using AllPrep DNA/RNA FFPE kit according to the manufacturer’s recommended protocol (Qiagen, Valencia, CA, USA). The Fusionplex RNA-sequencing assay was performed using a customized 115-gene panel covering a wide spectrum of cancer genes known for their involvement in gene fusions in neoplasia including, but not limited to, FN1, CSF1, FGFR1, and FGFR2 (ArcherDx, Inc. Boulder, CO). The methods for Fusionplex RNA-sequencing analysis have been described previously (10).

### RT-PCR and Sanger sequencing confirmation

For the positive fusions detected by targeted RNA-seq, RT-PCR and Sanger sequencing was performed to confirm the fusions and breakpoints at RNA level. cDNAs were synthesized by random priming with Fusionplex reagent kit (ArcherDX, Boulder, CO, USA), then were subjected to polymerase chain reaction (PCR) using FastStart Taq Polymerase (Roche Diagnostics, Indianapolis, IN, USA) with specific primers designed for each partner gene (Table S1). PCR products were cloned using TOPO TA Cloning Kit (Invitrogen, Carlsbad, CA, USA) and then were Sanger-sequenced using Eurofins Genomics Tube Sequencing service (Eurofins Genomics, Louisville, KY, USA).

## RESULTS

### Clinical and Pathologic Findings

Our series of 9 patients includes 4 women and 5 men with a mean age of 56.5 years (range 36 to 72 years). Anatomic locations include the temporomandibular joint and/or temporal bone in 5 cases (56%), hand/digits in 3 cases (33%), and foot in 1 case (11%). Tumor size, when available, ranged from 0.5 to 4.0 cm (mean 3.1 cm). All tumors were removed by surgical excision. Follow-up information was available for 2 cases, with a duration of 1 year each. No recurrence or metastasis was reported in any case. See Table 1 for complete clinicopathologic information.

**Table 1.** Clinicopathological features and molecular characterization of gene fusions.

| Number | Age | Sex | Location | Size (cm) | Recurrence | Metastasis | Fusion (5' exon::exon 3')<sup>a</sup> | Genomic coordinates [hg19] |
| --- | --- | --- | --- | --- | --- | --- | --- | --- |
| 1 | 70 | M | TMJ<sup>b</sup> | 4.0 cm | NA<sup>c</sup> | NA | FN1 → FGFR2 (E15::E7)<sup>d</sup> | chr2:216274286,chr10:123279683 |
| 2 | 66 | F | TMJ | 3.9 cm | No (1 year) | No | FN1 → FGFR2 (E42/ E38::E5)<sup>e</sup> | chr2:216232586/<br>chr2:216238045,chr10:123310973 |
| 3 | 69 | F | Foot | 3.8 cm | No (1 year) | No | FN1 → FGFR2 (E31::E5) | chr2:216246935,chr10:123310973 |
| 4 | 52 | F | Hand | 0.5 cm | NA | NA | FN1 → FGFR2 (E11::E5) | chr2:216285396,chr10:123310973 |
| 5 | 72 | F | TMJ | Unknown | NA | NA | FN1 → FGFR2 (E17::E5) | chr2:216272831,chr10:123310973 |
| 6 | 44 | M | TMJ | Unknown | NA | NA | FGFR1 → PLAG1 (E1::E3) | chr8:38325499,chr8:57083748 |
| 7 | 36 | M | 5th finger-palm | Unknown | NA | NA | FN1 → FGFR1 (E25::E9)<sup>f</sup> | chr2:216256355,chr8:38277253 |
| 8 | 51 | M | Thumb | Unknown | NA | NA | FN1 → MERTK (E24::E2) | chr2:216259251,chr2:112686697 |
| 9 | 49 | M | Temporal/external<br>auditory canal | Unknown | NA | NA | FN1 → TEK (E27::E13) | chr2:216251412,chr9:27202818 |

Note: <sup>a</sup>Exon numbers were based on the transcript numbers of genes used for *FN1* (NM\_002026.2), *FGFR2* (NM\_000141.4), *MERTK* (NM\_006343.2), *FGFR1* (NM\_015850.3), *TEK* (NM\_000459), and *PLAG1* (NM\_002655.2);
<sup>b</sup>TMJ, temporomandibular joint;
<sup>c</sup>NA, not available;
<sup>d</sup>Only breakpoints reported for *FN1-FGFR2* fusion (2);
<sup>e</sup>Two alternatively spliced variants detected in case 2;
<sup>f</sup>Breakpoints differ from what was reported for the *FN1-FGFR1* fusions detected in phosphaturic mesenchymal tumors (3, 12, 27, 28)

Histologically, all tumors showed multinodular architecture and chondroid to cartilaginous matrix (Fig. 1 A-B, cases 2 and 4) with increased cellularity towards the periphery of the nodules. The matrix frequently showed calcification that was coarse or grungy to lacey (chondroblastoma-like) (Fig. 1 C-D, cases 2 and 4). Remarkably, the grungy calcifications in case 5 were intensely basophilic, appeared crystalline (Fig. 2 A-B), and examination using polarized light revealed refractive rhomboid crystals consistent with calcium pyrophosphate dihydrate (CPPD) deposition(Fig. 2 C). The tumor cells within the chondroid/cartilage matrix were polygonal to stellate with abundant eosinophilic cytoplasm and eccentrically placed nuclei with small nucleoli, while the cells in fibrous septa were more often smaller and spindled with fibroblastic features (Fig. 3 A-C, cases 2, 3 and 8). While osteoclast-like giant cells were present in all cases and careful examination could often find at least focal areas resembling tenosynovial giant cell tumor (epithelioid to histiocytoid cells with eccentric nuclei and hemosiderin deposition), TGCT-like features were particularly prominent in case 9 (Fig. 4 A-B) which also had the least amount of chondroid stroma. For comparison, we tested a case in our archives previously classified as TGCT with focal chondroid metaplasia occurring in the TMJ (Figure 4 C-D), and that tumor was negative for gene fusions, including *FN1, FGFR1, FGFR2* and *CSF1*, using our targeted gene panel.

**Figure 1.**
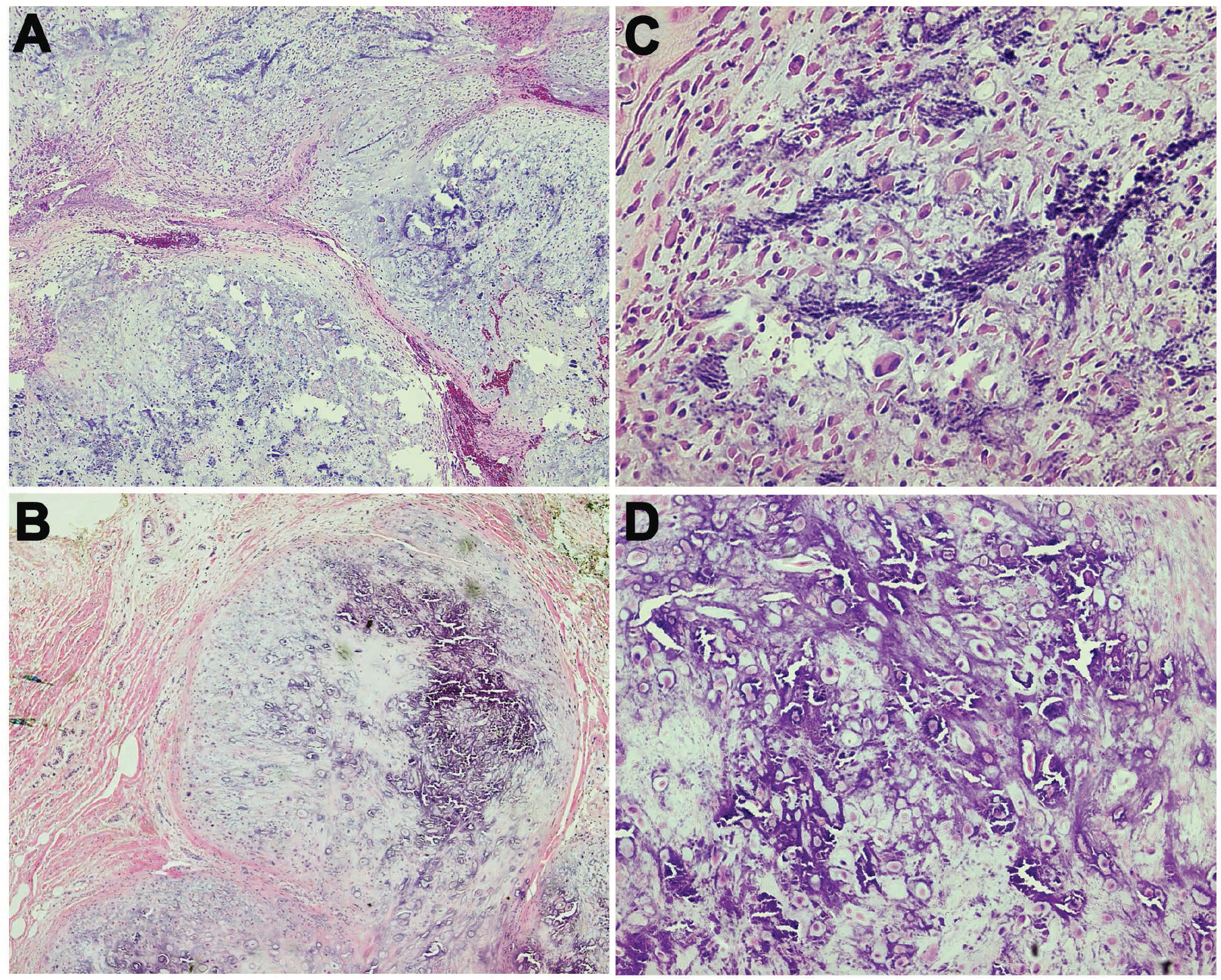
Major histologic features of soft tissue cartilage neoplasms in this study. (A-B) Characteristic lobular architecture in cases 2 (A) and 4 (B). (C-D) Grungy to lacey (chondroblastoma-like) calcifications in cases 2 (C) and 4 (D).

### Molecular Findings

By targeted RNA sequencing, an in-frame gene fusion was detected in all 9 cases. In 8 tumors, the fusion genes were comprised of *FN1* as the 5’ partner gene and various 3’partner genes including *FGFR2* in 5 cases (cases 1-5), *FGFR1* in one case (case 7) and novel partner genes *MERTK* and *TEK* in two cases (cases 8 and 9, respectively) (Table 1, Figure 2). An *FGFR1-PLAG1* fusion gene was detected in 1 tumor with *FGFR1* as the 5’ partner and *PLAG1* as the 3’ partner (case 6). All fusion transcripts were further verified by RT-PCR and Sanger sequencing (Figure S1).

**Figure 2.**
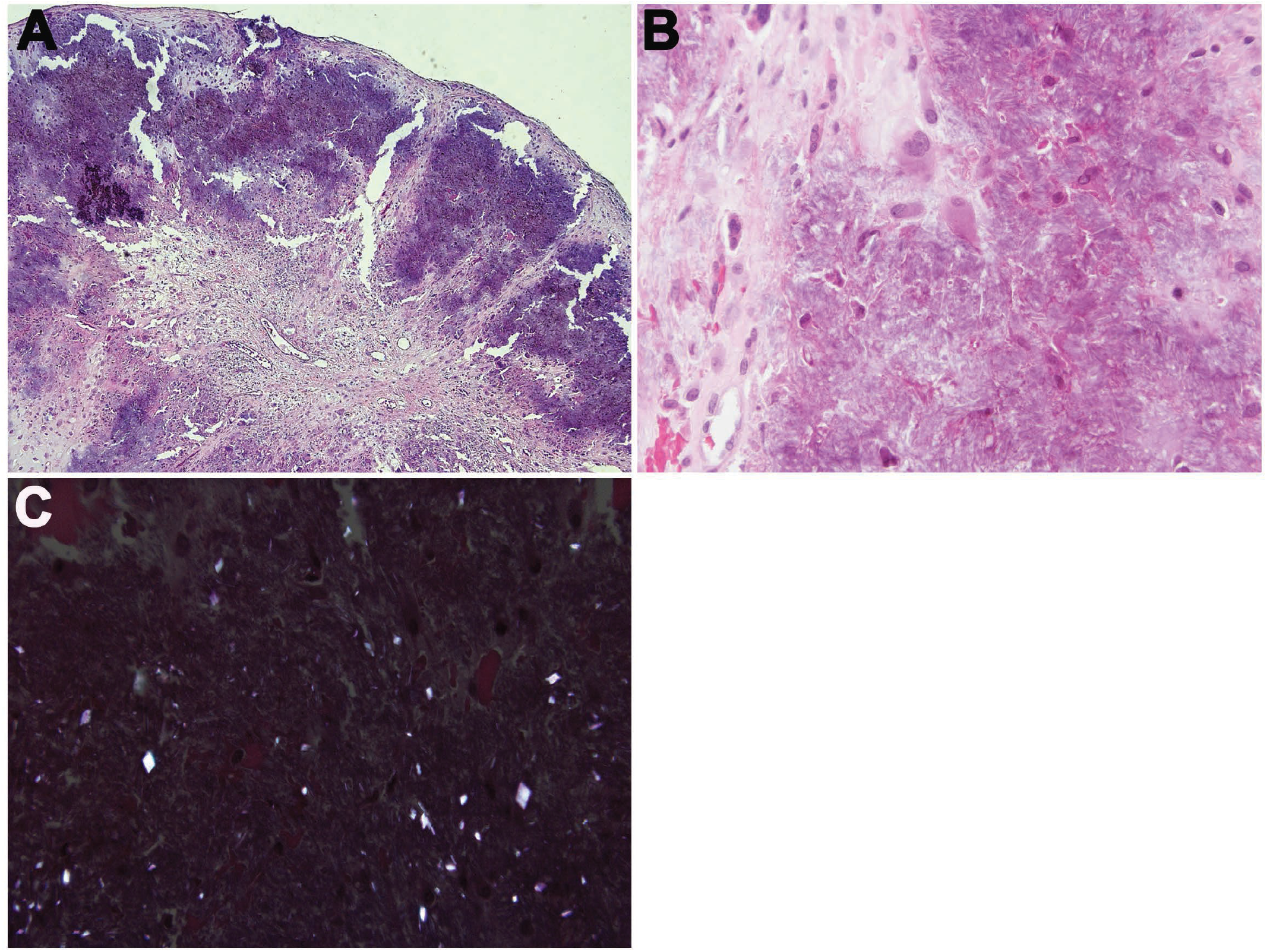
Case 5 with CPPD crystal deposition. (A-B) Case 5 with extensive basophilic grungy calcification at low power (A) and epithelioid to differentiated chondrocytes within crystalline deposits at high power (B). (C) Refractile rhomboid crystals when viewed using polarized light consistent with CPPD.

Gene fusions involved various breakpoints within the coding sequencing of the 5’ partner gene *FN1*, ranging from the 3’ end of exon 11 to exon 48, retaining signal peptide domain (SP), all FN type 1 domains (FN1 binding assembly domain) and FN type 2 domains, and up to sixteen FN type 3 domains (Figure 5). The various 3’partner genes of the fusion transcripts had breakpoints at the 5’ ends of exon 5 to exon 7 of *FRGR2* in 5 cases, exon 9 of *FGFR1*, exon 2 of *MERTK*, and exon 13 of *TEK* (Figure 5, Table 1), retaining trans-membrane domain (TM) and tyrosine kinase domain (TK) in all eight cases. However, up to two extracellular FGF-binding (Ig-like) domains were retained in the fusions, including two of three Ig-like domains (Ig2 and Ig3) of *FGFR2* in cases 2-5, Ig3 only of *FGFR2* in case 1, and Ig1 and Ig2 of *MERTK* in case 8, no Ig-like domain of *FGFR1* in case 7, and no Ig-like domain of *TEK* in case 9 (Figure 5, Table 1). Of note, two alternatively spliced fusion transcripts were detected in case 2. One tumor harbored an *FGFR1-PLAG1* fusion gene with breakpoints between exons 1 and 2 of *FGFR1* and between exons 2 and 3 of *PLAG1*.

**Figure 3.**
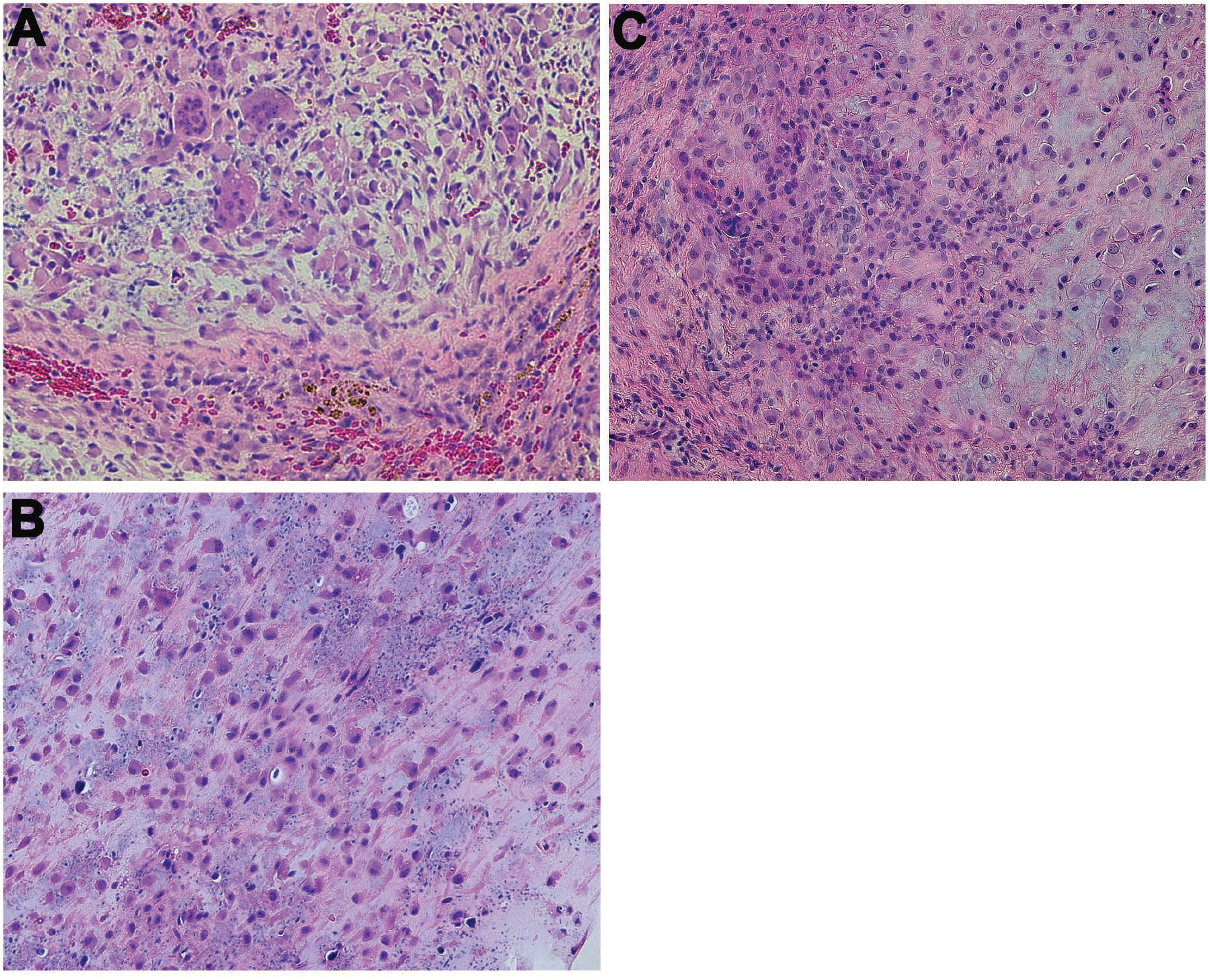
Cytologic features of the soft tissue cartilage neoplasms in this study. (A-C) Cells within chondroid matrix are polygonal to stellate with abundant eosinophilic cytoplasm and eccentrically placed nuclei with small nucleoli, while septae separating lobules contain spindled, fibroblastic cells. Osteoclast giant cells are also a frequent finding. (A) Case 2. (B) Case 3. (C) Case 8.

**Figure 4.**
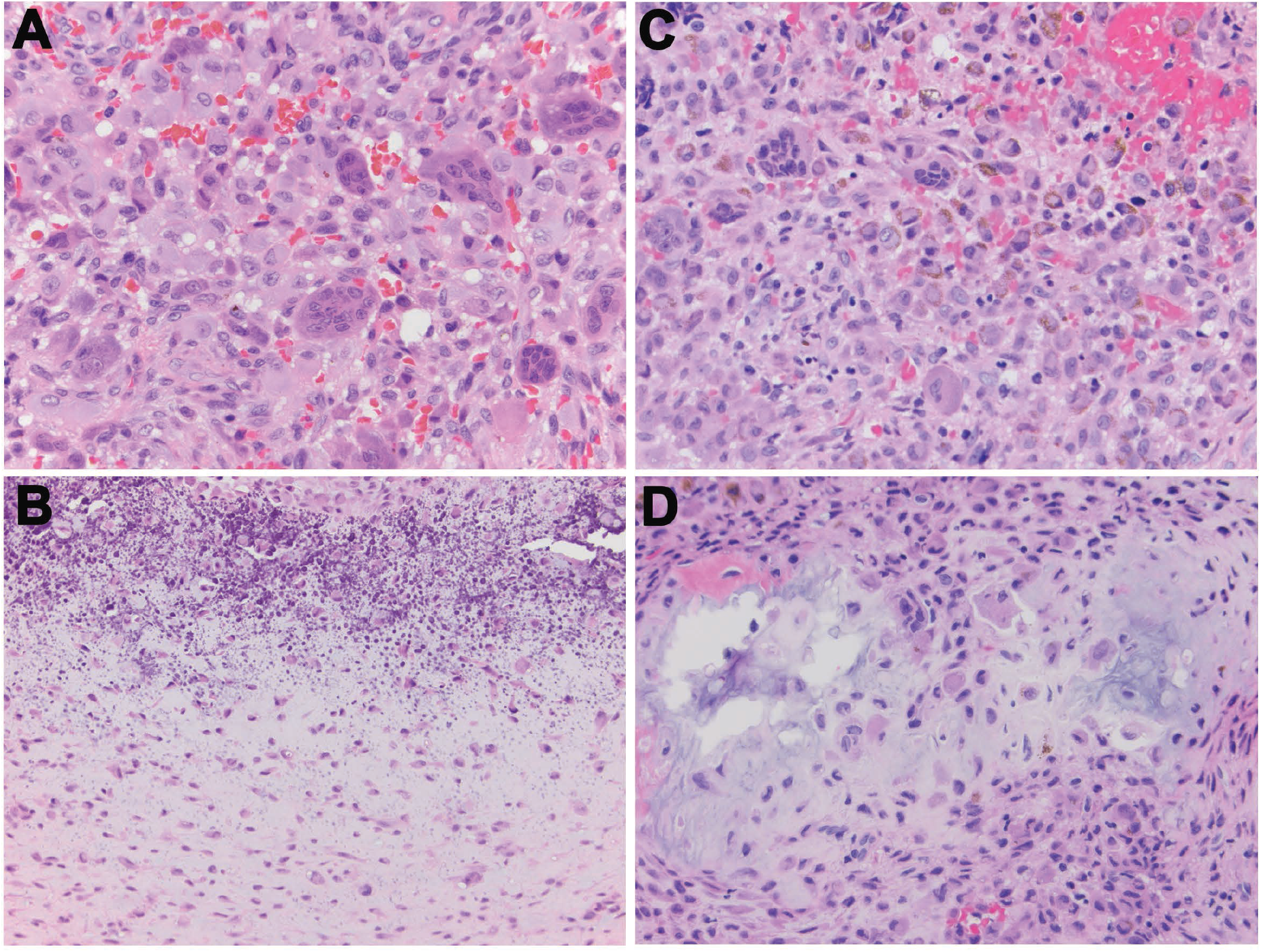
Soft tissue cartilage neoplasm in this study with features resembling tenosynovial giant cell tumor (TGCT). (A-B) Case 9 that predominantly shows the histologic features of epithelioid cells with eccentric nuclei admixed with osteoclast-like giant cells, resembling some features of a TGCT (A) as well as focal area of epithelioid to stellate cells in chondroid matrix with calcification (B). (C-D) Case of a TGCT occurring in the TMJ for comparison with case 9 showing mononuclear epithelioid/histiocytoid cells with eccentric nuclei and ring-like distribution of cytoplasmic hemosiderin as well as multinucleated giant cells (C). Focal area of chondroid metaplasia is present (D).

**Figure 5.**
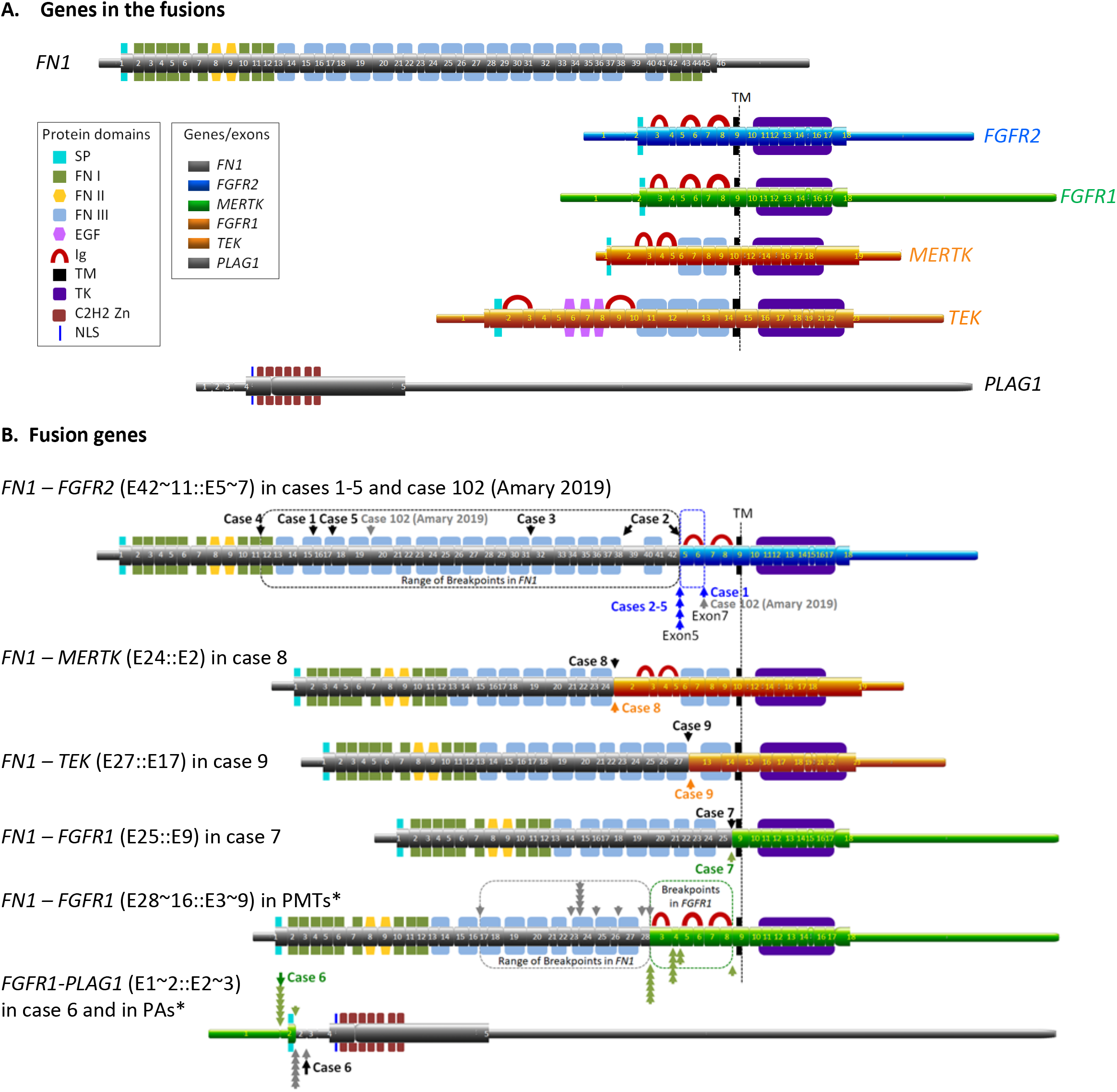
Schematic of chimeric transcripts and proteins resulting from gene fusions. (A). RNA transcripts with exon structures and related protein domains in the genes involved in the fusions. (B). Chimeric transcripts and proteins detected in the cohort of soft tissue chondroma of this study and related tumors in the literature, showing the retained exons and functional domains of the fusion genes of *FN1, FGFR2, FGFR1, MERTK*, and *PLAG1*. Case 102 is the only case with *FN1-FGFR2* characterized by RNA-seq previously (2). *PMTs = Phosphaturic Mesenchymal Tumors with *FN1-FGFR1* fusions and range of breakpoints reported in the literature (3,12,25,27,28). *PAs = pleomorphic adenoma of salivary gland origin with FGFR1-PLAG1 fusions and range of breakpoints reported in the literature (40). Untranslated regions (5’ UTR and 3’ UTR) are shown as narrow bars. Exons are shown as boxes with numbers. Protein domains are represented by shapes with keys shown in the box: SP = Signal Peptide, FN type 1 domain, FN type 2 domain, FN type 3 domain, EGF=EGF-like domain, Ig=Immunoglobulin-Like loop domain, TM = Transmembrane domain, and TK = Tyrosine Kinase, C2H2 Zn =C2H2 Zn finger, and NLS=Nuclear localization signal.

## DISCUSSION

In this study we characterize the presence of *FN1* fusion with genes encoding receptor tyrosine kinases in 8 calcified chondroid mesenchymal neoplasms as well as *FGFR1-PLAG1* in one tumor (Table 1). Gene fusions involving *FN1-FGFR2* or *FN1-FGFR1* have been recently described in a subset of soft tissue chondromas, which were mostly demonstrated indirectly using break-apart FISH probes for *FN1, FGFR1*, and/or *FGFR2* (2). To our knowledge, only one case of soft tissue chondroma with *FN1-FGFR2* has been characterized by RNA sequencing and showed a fusion between exons 1-19 of *FN1* and exons 7-17 of *FGFR2* (2). The breakpoints in *FN1* and *FGFR2* reported in this case are in the range of breakpoints defined in the five tumors harboring *FN1-FGFR2* fusions in our series (Figure 5B). We also describe two novel gene fusions, *FN1-MERTK* and *FN1-TEK* (Table 1, Figure 5). *FN1* encodes fibronectin-1, a glycoprotein present in a dimeric form in plasma and in dimeric/multimeric forms at the cell surface and in extracellular matrix, which participates in cell adhesion and migration processes (11). All fusion partners of *FN1* detected in our series encode for receptor tyrosine kinases; these include *FGFR2* (fibroblast growth factor receptor 2), *FGFR1* and *MERTK* (MER proto-oncogene, tyrosine kinase). These fusions result in the in-frame fusion of the N-terminal region of FN1 to the intact transmembrane and tyrosine kinase domains of the receptor tyrosine kinase, and as a result, may promote dimerization of the fused receptor through the fibronectin domain and thereby lead to aberrant signal activation as previously hypothesized (3). The presence of fusions involving these potentially targetable receptor tyrosine kinases suggests potential alternative therapeutic avenues for treating such tumors if needed.

Fusions of *FN1* to other receptor tyrosine kinases have also been detected in a variety of neoplasms other than soft tissue chondroma, including *FN1-FGFR1* and *FN1-FGF1* in phosphaturic mesenchymal tumor (3,12), *FN1-EGF* in lipofibromatosis and calcifying aponeurotic fibroma (1,13), *FN1-AVCR2A* in synovial chondromatosis (2), *FN1-ALK* in gastrointestinal leiomyoma and inflammatory myofibroblastic tumor (14-16), *FN1-1GF1R* in ALK-negative inflammatory myofibroblastic tumor (17), and *FN1-ROS1* in infantile inflammatory myofibroblastic tumors (18).

Soft tissue chondromas in general are characterized by nodular/lobular growth of well-differentiated chondrocytes. However, the chondroblastoma-like variant of soft tissue chondroma as described by Cates et al. (2001) also shows cellular foci of epithelioid chondrocytes of varying size admixed with osteoclast-like giant cells within a variable amount of chondroid matrix that is frequently accompanied by lace-like calcification (19). While most cases of soft tissue chondroma have been documented to occur in the extremities, lesions involving the parotid region and skull base has been reported (20). The cases in our series, particularly the ones with *FN1-FGFR1, FN1-FGFR2* and *FN1-MERTK* fusions, show significant morphologic overlap with previous morphologic descriptions of the chondroblastoma-like variant of soft tissue chondroma; however, we also describe TGCT-like features as well as CPPD crystal deposition, expanding the morphologic spectrum of this group of entities. We also report a larger proportion affecting the temporomandibular joint (TMJ) region. We therefore believe that this group of neoplasms may not be best classified as simply a variant of soft tissue chondroma and have chosen to apply the term “calcified chondroid mesenchymal neoplasm” for the purposes of this study.

A few of the cases in our series warrant special consideration (see a summary of differential diagnosis in table 2). One tumor involving the TMJ and harboring *FN1-FGFR2* (case 5) showed extensive deposition of calcium pyrophosphate dehydrate (CPPD) crystals. Tophaceous pseudogout is a mass-forming (tumoral) deposition of CPPD crystals that has been described in a number of locations near joints including the TMJ and some show erosion into the skull base (7,21,22). Cellular infiltrates resembling foreign body type giant cell reaction are typically present, and cartilage/chondroid tissue (supposedly metaplastic) may also be seen in a significant subset of examples (7,22). While only one fusion-positive case in our series contained CPPD deposition, this observation calls into question whether tumoral (massive) tophaceous pseudogout, or at least a subset of cases, actually represent neoplasms harboring recurrent gene fusions.

**Table 2.**
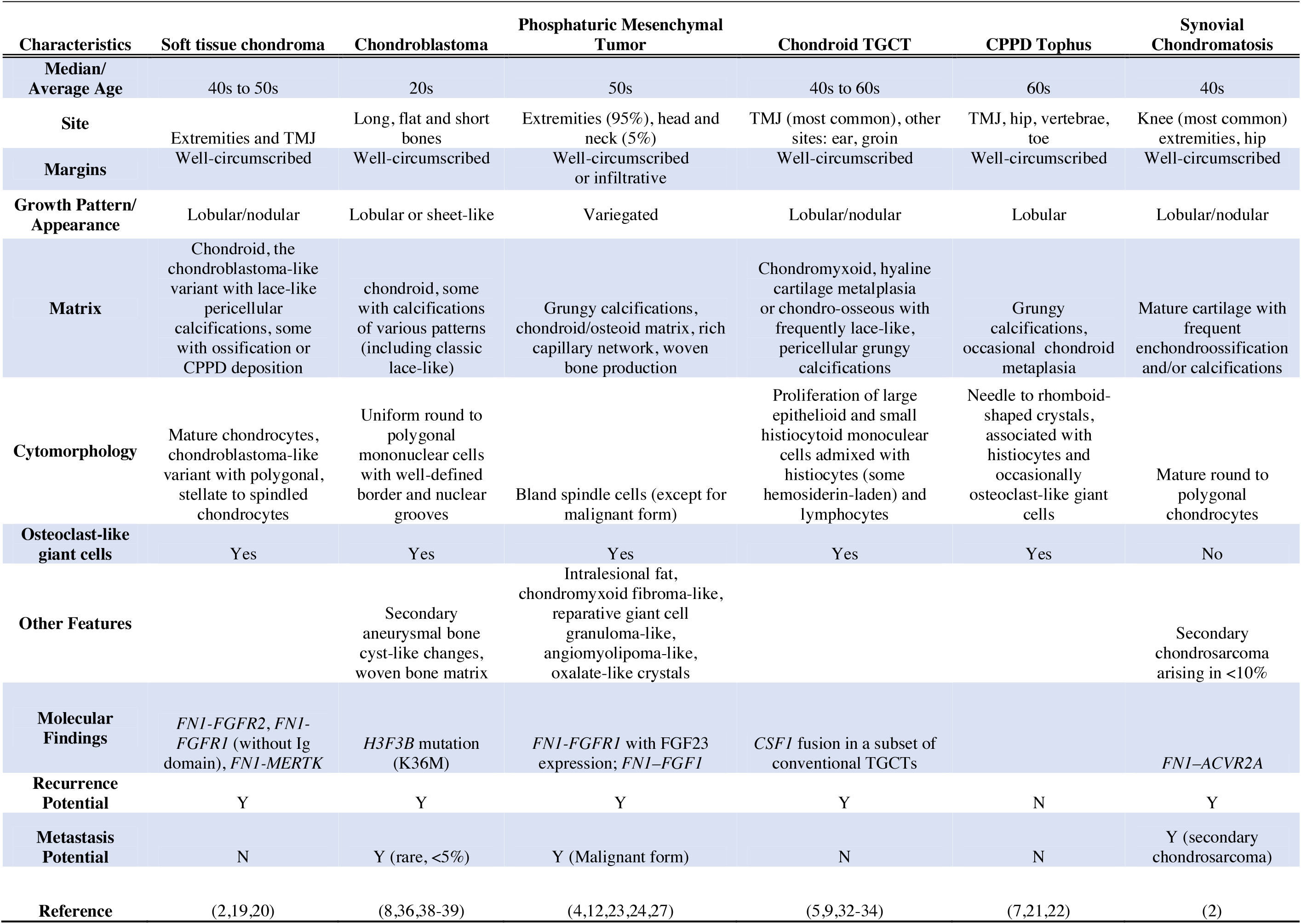
Summary of differential diagnosis.

For the one tumor in our series harboring an *FN1-FGFR1* fusion gene (Figure 5B, case 7), we considered the possibility of phosphaturic mesenchymal tumor (PMT). PMTs can take on a wide range of histologic appearances, but in general are characterized having a highly vascular stroma (some with hemangiopericytoma-like pattern) with bland spindled to stellate neoplastic cells and amorphous basophilic chondro-osseous matrix with grungy calcification (4,23,24). A significant subset of PMT was also shown to harbor *FN1-FGFR1* fusions and are frequently associated with hypophosphatemia and tumor-induced osteomalacia secondary to paraneoplastic secretion of the fibroblast growth factor 23 (FGF23) (3,12). Examples of PMT without apparent tumor-induced osteomalacia, however, have also been described (24) as well as fusion-negative cases that frequently overexpress α-klotho, a transmembrane enzyme that acts as an FGF23 activator (25). α-Klotho interacts directly with FGFR1 and forms a high-affinity binding site at Ig3 domain for FGF23 (26). None of the cases in our series, including the one with *FN1-FGFR1*, exhibited the highly vascular stroma or spindle cell proliferation typical of PMT and none of these patients had any known osteomalacia. Given the absence of phosphaturia and definitive histologic features of PMT, this case is best classified as a chondroblastoma-like soft tissue chondroma. Furthermore, unlike most of *FN1-FGFR1* fusions characterized by sequencing in PMT which retained mostly all 3 or 2 of Ig domains of *FGFR1* (3,12,27,28), *FN1-FGFR1* fusion in case 7 did not contain any Ig domains of *FGFR1* (Figure 5B). Without the Ig3 domain, the *FN1-FGFR1* fusion protein could not interact with FGF23. The recent description of *FN1-FGFR1* fusions in three cases of soft tissue chondroma were indirectly demonstrated using break-apart FISH probes for *FN1* and *FGFR1* without the knowledge of protein domains retained (2). It would be informative to assess whether these three cases retain any Ig domain. The lack of significant levels of FGF23 in these three cases of soft tissue chondromas suggest a potential lack of Ig domains in their fusions. In all, our study has demonstrated the effectiveness of RNA-seq as a tool for identifying functional domains retained in gene fusions and in turn facilitating correlation of molecular findings with clinicopathological features and tumor classification.

We detected an *FGFR1-PLAG1* in one case involving the TMJ. *FGFR1-PLAG1* fusions have been previously detected in pleomorphic adenoma (PA) of salivary gland origin and, in such cases, the 5’-portion of *FGFR1*, excluding the tyrosine kinase domain, is fused to the entire *PLAG1* coding sequence (29,30). This fusion gene arrangement is the same as that found in case 6 of our series. While PA is principally defined histologically by the proliferation of ductal and myoepithelial cells within hyalinized to myxoid stroma, cartilaginous components may also be seen and can be quite extensive (31). Our case was entirely composed of lobular cartilaginous tissue with variable, often grungy, calcification without any identifiable ductal or myoepithelial population and therefore histologically appeared essentially indistinguishable from the other chondroblastoma-like soft tissue chondromas in our series. In such an archived case, excluding the possibility of a PA with extensive cartilaginous components in which ductal/myoepithelial components were either not present or not sampled is problematic; however, we found no morphologic features to classify it as PA. We must also consider the possibility that *FGFR1-PLAG1* may represent an alternate fusion gene found in non-PA soft tissue cartilaginous neoplasms.

Chondroid tenosynovial giant cell tumor (TGCT) is a rare tumor with a predilection for the TMJ region and skull base that often demonstrates locally aggressive growth including bone destruction (5,6,32-34). Histologically, chondroid TGCT shows features of conventional TGCT, such as sheet-like proliferation of large epithelioid to histiocytoid mononuclear cells, some with hemosiderin deposition often in a ring like deposition around the cytoplasm, and multinucleated giant cells; however, geographic or nodular areas of metaplastic chondroid matrix are also present and frequently associated with grungy, lace-like calcifications. While the majority of conventional TGCTs have been shown to harbor *CSF1* fusions (35), this disease-specific genetic event has not been demonstrated in chondroid TCGT of the TMJ. The case with *FN1-TEK* fusion in our study shows prominent morphologic features resembling TGCT with focal areas of chondroid matrix and calcifications as seen in the other cases in the series. This finding raises the possibility that lesions previously classified as chondroid TGCT may actually fall into this group of calcified chondroid mesenchymal neoplasms with *FN1* gene fusions. For comparison, we tested another case from our archives previously classified as TGCT with chondroid metaplasia occurring in the TMJ (Figure 4 C-D), and that tumor tested negative for gene fusions, including *FN1, FGFR1, FGFR2* and *CSF1*, using our targeted gene panel. It remains to be determined whether any other tumors previously described as chondroid TGCT harbor recurrent gene fusions including an *FN1*-tyrosine kinase receptor fusion.

Additional differential diagnostic considerations include chondroblastoma and synovial chondromatosis. Chondroblastoma most often occurs in the epiphyses of long bones but may also affect the craniofacial skeleton. Histologically, it is characterized by sheets of uniform round to polygonal mononuclear cells with distinct cell borders and grooved nuclei with scattered islands of cartilaginous differentiation often showing lacey (“chicken-wire”) calcification (36). While some of our cases show a lacey calcification pattern within the cartilaginous elements, our tumors generally lack the sheet-like cellularity and characteristic cytomorphology of chondroblastoma. Lacey (chondroblastoma-like) calcifications have also been previously described in chondroblastoma-like soft tissue chondroma (19,20,37). In addition, *H3F3B* K36M mutation has been detected in greater than 90% of chondroblastomas and immunohistochemical staining using a mutation specific antibody has been shown to be highly sensitive and specific (38,39). Immunohistochemical staining for the K36M mutant was negative in one case with *FN1-FGFR2* fusion (Case 2). Synovial chondromatosis is characterized by a strikingly nodular proliferation of fairly uniform, mature, hyaline, cartilaginous tissue, often with distinct clustering of chondrocytes, which involves the joint synovium, tendon sheath, or bursa. A subset of cases harbors *FN1-ACVR2A* fusion and this finding has been described as a distinguishing feature of synovial chondromatosis from soft tissue chondroma (2). While the tumors in our series do show lobulated growth of cartilaginous tissue, they lack the distinct clustering of chondrocytes and are negative for the *FN1-ACVR2* fusion gene. Overall, our findings indicate that the tumors in our series represent distinct entities from both chondroblastoma and synovial chondromatosis.

In summary, our study presents 9 examples of “calcified chondroid mesenchymal neoplasm,” 8 of which harbor FN1-receptor tyrosine kinase fusions showing a predilection for the TMJ region as well as the extremities. The *FN1* fusions with receptor tyrosine kinase genes include *FN1-FGFR2, FN1-FGFR1* (without Ig3 domain), as well as the novel fusions *FN1-MERTK* and *FN1-TEK*. Unlike the *FN1-FGFR1* fusions found in PMT, the *FN1-FGFR1* fusion detected in our series did not retain the Ig3 domain of FGF23 binding site. While most cases in our series (particularly the ones with *FN1-FGFR1, FN1-FGFR2* and *FN1-MERTK1* fusions) show morphologic overlap with chondroblastoma-like soft tissue chondroma, we also report the novel findings of extensive CPPD crystal deposition in one case and extensive morphologic features resembling TGCT, expanding the morphologic spectrum of tumors with *FN1*-receptor tyrosine kinase fusions beyond those previously described for soft tissue chondroma. Our findings also raise the question of whether some lesions previously classified as tophaceous pseudogout and chondroid TGCT may actually represent neoplasms harboring recurrent *FN1* gene fusions. The presence of *FGFR1-PLAG1* in one of our cases suggests that pleomorphic adenoma of salivary gland origin should be considered in the differential diagnosis of cartilage lesions affecting the craniofacial region. While most of these tumors are amenable to surgical excision, the structure of these fusion genes indicates that therapeutic targeting of the receptor tyrosine kinases may be a promising alternative treatment avenue if needed.

## Data Availability

All sequencing data will be made available upon manuscript acceptance at a journal.

## ACKNOWLEDGMENTS

This project is supported by intra-departmental funding.

## DISCLOSURE/CONFLICT OF INTEREST

The authors have no conflict of interest to declare.

## Notes

### Competing Interest Statement

The authors have declared no competing interest.

### Funding Statement

The work is supported by intra-departmental fund only. No external funding was used. The authors did not receive payment or services from a third party for any aspect of the submitted work.

### Author Declarations

This project was approved by the Institutional Review Board at the University of Washington.

## REFERENCES

1. Puls F, Hofvander J, Magnusson L, Nilsson J, Haywood E, Sumathi VP, et al. FN1-EGF gene fusions are recurrent in calcifying aponeurotic fibroma. J Pathol. 2016 Mar;23 8(4):5 02-7.

2. Amary F, Perez-Casanova L, Ye H, Cottone L, Strobl A-C, Cool P, et al. Synovial chondromatosis and soft tissue chondroma: extraosseous cartilaginous tumor defined by FN1 gene rearrangement. Mod Pathol. 2019 Dec;32(12):1762–71.

3. Lee J-C, Jeng Y-M, Su S-Y, Wu C-T, Tsai K-S, Lee C-H, et al. Identification of a novel FN1-FGFR1 genetic fusion as a frequent event in phosphaturic mesenchymal tumour. J Pathol. 2015 Mar;235(4):539–45.

4. Folpe AL, Fanburg-Smith JC, Billings SD, Bisceglia M, Bertoni F, Cho JY, et al. Most osteomalacia-associated mesenchymal tumors are a single histopathologic entity: an analysis of 32 cases and a comprehensive review of the literature. Am J Surg Pathol. 2004 Jan;28(1):1–30.

5. Hoch BL, Garcia RA, Smalberger GJ. Chondroid tenosynovial giant cell tumor: a clinicopathological and immunohistochemical analysis of 5 new cases. Int J Surg Pathol. 2011 Apr;19(2):180–7.

6. Oda Y, Izumi T, Harimaya K, Segawa Y, Ishihara S, Komune S, et al. Pigmented villonodular synovitis with chondroid metaplasia, resembling chondroblastoma of the bone: a report of three cases. Mod Pathol. 2007 May;20(5): 545–51.

7. Ishida T, Dorfman HD, Bullough PG. Tophaceous pseudogout (tumoral calcium pyrophosphate dihydrate crystal deposition disease). Hum Pathol. 1995 Jun;26(6):587–93.

8. Konishi E, Nakashima Y, Mano M, Tomita Y, Kubo T, Araki N, et al. Chondroblastoma of extra-craniofacial bones: Clinicopathological analyses of 103 cases. Pathol Int. 2017 0ct;67(10):495–502.

9. Wang J-G, Liu J, He B, Gao L, Zhang L, Liu J. Diffuse Tenosynovial Giant Cell Tumor Around the Temporomandibular Joint: An Entity With Special Radiologic and Pathologic Features. J Oral Maxillofac Surg. 2019 May;77(5):1022.e1–1022.e39.

10. Tretiakova MS, Wang W, Wu Y, Tykodi SS, True L, Liu YJ. Gene fusion analysis in renal cell carcinoma by FusionPlex RNA-sequencing and correlations of molecular findings with clinicopathological features. Genes Chromosomes Cancer. 2019 Aug 10;

11. Zollinger AJ, Smith ML. Fibronectin, the extracellular glue. Matrix Biol. 2017;60-61:27–37.

12. Lee J-C, Su S-Y, Changou CA, Yang R-S, Tsai K-S, Collins MT, et al. Characterization of FN1-FGFR1 and novel FN1-FGF1 fusion genes in a large series of phosphaturic mesenchymal tumors. Mod Pathol. 2016;29(11):1335–46.

13. Al-Ibraheemi A, Folpe AL, Perez-Atayde AR, Perry K, Hofvander J, Arbajian E, et al. Aberrant receptor tyrosine kinase signaling in lipofibromatosis: a clinicopathological and molecular genetic study of 20 cases. Mod Pathol. 2019;32(3):423–34.

14. Haimes JD, Stewart CJR, Kudlow BA, Culver BP, Meng B, Koay E, et al. Uterine Inflammatory Myofibroblastic Tumors Frequently Harbor ALK Fusions With IGFBP5 and THBS1. Am J Surg Pathol. 2017 Jun;41(6):773–80.

15. Panagopoulos I, Gorunova L, Lund-Iversen M, Lobmaier I, Bjerkehagen B, Heim S. Recurrent fusion of the genes FN1 and ALK in gastrointestinal leiomyomas. Mod Pathol. 2016;29(11): 1415–23.

16. Ouchi K, Miyachi M, Tsuma Y, Tsuchiya K, Iehara T, Konishi E, et al. FN1: a novel fusion partner of ALK in an inflammatory myofibroblastic tumor. Pediatr Blood Cancer. 2015 May;62(5):909–11.

17. Piarulli G, Puls F, Wängberg B, Fagman H, Hansson M, Nilsson J, et al. Gene fusion involving the insulin-like growth factor 1 receptor in an ALK-negative inflammatory myofibroblastic tumour. Histopathology. 2019 Jun;74(7):1098–102.

18. Lopez-Nunez O, John I, Panasiti RN, Ranganathan S, Santoro L, Grélaud D, et al. Infantile inflammatory myofibroblastic tumors: clinicopathological and molecular characterization of 12 cases. Mod Pathol. 2020;33(4):576–90.

19. Cates JM, Rosenberg AE, O’Connell JX, Nielsen GP. Chondroblastoma-like chondroma of soft tissue: an underrecognized variant and its differential diagnosis. Am J Surg Pathol. 2001 May;25(5):661–6.

20. Raparia K, Lin JW, Donovan D, Vrabec JT, Zhai QJ, Ayala AA, et al. Chondroblastoma-like chondroma of soft tissue: report of the first case in the base of skull. Ann Diagn Pathol. 2013 Jun;17(3):298–301.

21. Houghton D, Munir N, Triantafyllou A, Begley A. Tophaceous pseudogout of the temporomandibular joint with erosion into the middle cranial fossa. Int J Oral Maxillofac Surg. 2020 Apr 8;

22. Kurihara K, Mizuseki K, Saiki T, Wakisaka H, Maruyama S, Sonobe J. Tophaceous pseudogout of the temporomandibular joint: report of a case. Pathol Int. 1997 Aug;47(8):578–80.

23. Agaimy A, Michal M, Chiosea S, Petersson F, Hadravsky L, Kristiansen G, et al. Phosphaturic Mesenchymal Tumors: Clinicopathologic, Immunohistochemical and Molecular Analysis of 22 Cases Expanding their Morphologic and Immunophenotypic Spectrum. Am J Surg Pathol. 2017 Oct;41(10):1371–80.

24. Sent-Doux KN, Mackinnon C, Lee J-C, Folpe AL, Habeeb O. Phosphaturic mesenchymal tumor without osteomalacia: additional confirmation of the “nonphosphaturic” variant, with emphasis on the roles of FGF23 chromogenic in situ hybridization and FN1-FGFR1 fluorescence in situ hybridization. Hum Pathol. 2018;80:94–8.

25. Lee C-H, Su S-Y, Sittampalam K, Chen PC-H, Petersson F, Kao Y-C, et al. Frequent overexpression of klotho in fusion-negative phosphaturic mesenchymal tumors with tumorigenic implications. Mod Pathol. 2019 Dec 2;

26. Chen G, Liu Y, Goetz R, Fu L, Jayaraman S, Hu M-C, et al. α-Klotho is a non-enzymatic molecular scaffold for FGF23 hormone signalling. Nature. 2018 25;553(7689):461–6.

27. Yamada Y, Kinoshita I, Kenichi K, Yamamoto H, Iwasaki T, Otsuka H, et al. Histopathological and genetic review of phosphaturic mesenchymal tumours, mixed connective tissue variant. Histopathology. 2018 Feb;72(3):460–71.

28. Saba KH, Cornmark L, Rissler M, Fioretos T, Äström K, Haglund F, et al. Genetic profiling of a chondroblastoma-like osteosarcoma/malignant phosphaturic mesenchymal tumor of bone reveals a homozygous deletion of CDKN2A, intragenic deletion of DMD, and a targetable FN1-FGFR1 gene fusion. Genes Chromosomes Cancer. 2019;58(10):731–6.

29. Katabi N, Xu B, Jungbluth AA, Zhang L, Shao SY, Lane J, et al. PLAG1 immunohistochemistry is a sensitive marker for pleomorphic adenoma: a comparative study with PLAG1 genetic abnormalities. Histopathology. 2018 Jan;72(2):285–93.

30. Persson F, Winnes M, Andrén Y, Wedell B, Dahlenfors R, Asp J, et al. High-resolution array CGH analysis of salivary gland tumors reveals fusion and amplification of the FGFR1 and PLAG1 genes in ring chromosomes. Oncogene. 2008 May 8;27(21):3072–80.

31. Klijanienko J, Servois V, Jammet P, Validire P, Pouillart P, Vielh P, et al. Pleomorphic adenoma. Am J Surg Pathol. 1998 Jun;22(6):772–3.

32. Fisher M, Biddinger P, Folpe AL, McKinnon B. Chondroid tenosynovial giant cell tumor of the temporal bone. Otol Neurotol. 2013 Aug;34(6):e49–50.

33. Carlson ML, Osetinsky LM, Alon EE, Inwards CY, Lane JI, Moore EJ. Tenosynovial giant cell tumors of the temporomandibular joint and lateral skull base: Review of 11 cases. Laryngoscope. 2017;127(10):2340–6.

34. Anbinder AL, Geraldo BMC, Guimaräes R, Pereira DL, Almeida OP de, Carvalho YR. Chondroid Tenosynovial Giant Cell Tumor of the Temporomandibular Joint: A Rare Case Report. Braz Dent J. 2017 Oct;28(5):647–52.

35. West RB, Rubin BP, Miller MA, Subramanian S, Kaygusuz G, Montgomery K, et al. A landscape effect in tenosynovial giant-cell tumor from activation of CSF1 expression by a translocation in a minority of tumor cells. Proc Natl Acad Sci USA. 2006 Jan 17;103(3):690–5.

36. Chen W, DiFrancesco LM. Chondroblastoma: An Update. Arch Pathol Lab Med. 2017 Jun;141(6):867–71.

37. Kuprys TK, Bindra R, Borys D, Nystrom LM. Chondroblastoma-like chondroma of the hand: case report. J Hand Surg Am. 2014 May;39(5):933–6.

38. Behjati S, Tarpey PS, Presneau N, Scheipl S, Pillay N, Van Loo P, et al. Distinct H3F3A and H3F3B driver mutations define chondroblastoma and giant cell tumor of bone. Nat Genet. 2013 Dec;45(12):1479–82.

39. Amary MF, Berisha F, Mozela R, Gibbons R, Guttridge A, O’Donnell P, et al. The H3F3 K36M mutant antibody is a sensitive and specific marker for the diagnosis of chondroblastoma. Histopathology. 2016 Jul;69(1):121–7.

40. Dalin MG, Katabi N, Persson M, Lee K-W, Makarov V, Desrichard A, et al. Multi-dimensional genomic analysis of myoepithelial carcinoma identifies prevalent oncogenic gene fusions. Nat Commun. 2017 30;8(1):1197.

